# Defecation During Hospitalization for Acute Coronary Syndrome and Future Cardiovascular Events

**DOI:** 10.1101/2023.12.07.23299700

**Authors:** Yasushi Matsuzawa, Kenichi Tsujita, Masaomi Gohbara, Masanobu Ishii, Taishi Nakamura, Hisaya Kondo, Tomohiro Yoshii, Ryusuke Sekii, Jin Kirigaya, Kengo Terasaka, Hidefumi Nakahashi, Eiichi Akiyama, Masaaki Konishi, Toshihiro Yamada, Yuichiro Arima, Shinsuke Hanatani, Seiji Takashio, Hiroki Usuku, Eiichiro Yamamoto, Masami Kosuge, Kazuo Kimura, Kiyoshi Hibi

## Abstract

**Background:** Gut function is vital for human health, and defecation frequency can serve as a fundamental marker including various abnormal patterns. However, the link between a defecation pattern and future adverse events in patients with acute coronary syndrome (ACS) remains uncertain. The aim of this study was to investigate the association between defecation patterns during hospitalization and future cardiovascular events in patients with ACS.

**Methods:** This two-center retrospective observational cohort study included 1949 patients hospitalized for ACS between 2012 and 2019. For a comprehensive assessment of defecation in a general ward, we examined three indicators: “frequency of non-defecation days,” “consecutive non-defecation days,” and “maximum daily defecation frequency,” in addition to “average daily defecation frequency”. Patients were divided according to Youden index-derived cutoff values of each defecation frequency indicators. The primary outcome was a composite of all-cause mortality, myocardial infarction, ischemic stroke, and hemorrhagic stroke.

**Results:** During the follow-up period (median, 48 months; IQR 30–74 months), 405 of 1949 (20.8%) patients developed the primary outcome. In total, 229 patients died (96 cardiovascular deaths and 133 non-cardiovascular deaths), and there were 142, 57, 28, and 113 patients with non-fatal myocardial infarction, nonfatal ischemic stroke, nonfatal hemorrhagic stroke, and hospitalized due to heart failure, respectively. High “frequency of non-defecation days” (≥33.5%) (hazard ratio [HR], 1.507; 95% confidence interval [95%CI], 1.191–1.907; *P*=0.0006) and high “maximum daily defecations frequency” (≥ 5 times in a single day) (HR, 1.670; 95%CI, 1.203–2.317; *P*=0.002) were associated with an increased risk of future cardiovascular events in the multivariate models. High “frequency of non-defecation days” and high “maximum daily defecations frequency” exhibit distinct characteristics: the former was associated with long-term cardiovascular mortality, all-cause mortality, myocardial infarction, and cerebral hemorrhage, while the latter was linked to cancer-related mortality, non-cardiovascular mortality, cerebral infarction, and heart failure.

**Conclusions:** Among patients with ACS who survived to discharge, abnormal defecation patterns as assessed by increased frequency of non-defecation days and high-frequency defecations within a single day, were independently associated with future cardiovascular events.

**Clinical Perspective:** *What Is New?:*

- This is the first study to investigate defecation patterns during the acute phase in patients with ACS and utilize reliable records of defecation during hospitalization rather than relying on survey-based assessments.
- In patients with ACS, both increased frequency of non-defecation days and high-frequency defecations within a single day were independently associated with an increased risk of future cardiovascular events, controlling for various confounding factors including age, severity of ACS, medications, and dietary intake.

*What Are the Clinical Implications?:*

- Abnormal defecation patterns, such as frequent non-defecation days and high-frequency defecations within a single day, serve as indicators of “gut frailty.”
- The presence of these abnormal defecation patterns may suggest a residual risk in patients post-ACS. Additional research is essential to explore the underlying mechanisms and potential therapeutic interventions.

## Introduction

Patients who survive acute coronary syndrome (ACS) are at a significantly elevated risk of recurrent ACS, stroke, cardiovascular mortality, and overall mortality, despite the implementation of secondary prevention measures.^1^

The statement “all disease begins in the gut,” attributed to the ancient Greek physician Hippocrates nearly 2500 years ago, has gained renewed relevance recently. The risk of cardiovascular events increases during straining in patients with constipation,^2^ but the gut plays a much more significant role in various aspects. Abnormal gut function may potentially contribute to cardiovascular diseases through mechanisms, including gut microbial dysbiosis, impairment of intestinal barrier function (commonly referred to as “Leaky gut syndrome”), systemic inflammation, autonomic nervous system dysregulation, and reduced secretion of cardioprotective incretins, such as glucagon-like peptide-1 and glucose-dependent insulinotropic polypeptide.^3–5^

Previous studies have reported an association between chronic constipation and cardiovascular mortality as well as non-fatal cardiovascular events.^6–8^ During acute illness, many factors, including sympathetic nervous system activation, systemic inflammatory response, reduced physical activity, and medication usage, can collectively influence gut function. Physical frailty during hospitalization for acute myocardial infarction and subsequent hospital-acquired disability can have a long-term adverse impact on prognosis.^9^ Nevertheless, the relationship between defecation frequency during acute illness and subsequent cardiovascular events remains unexplored. Furthermore, there is no established quantitative method for assessing defecation frequency during hospitalization. There can be several patterns of abnormal defecation, such as an increased number of non-defecation days (i.e., defecation every other day), consecutive non-defecation days, and high-frequency defecations within a single day. Therefore, we assessed the defecation patterns using three indicators: “frequency of non-defecation days,” “consecutive non-defecation days,” and “maximum daily defecation frequency,” in addition to “average daily defecation frequency” which is used in the definition of chronic constipation.

We hypothesized that defecation frequency, an indicator of “gut frailty,”^10^ is a residual risk factor in patients with ACS. Our primary aim was to examine the relationship between defecation frequency during a general ward stay after intensive care and the occurrence of future cardiovascular events in patients with ACS.

## Methods

### Study design and population

We conducted a retrospective, observational cohort study and enrolled consecutive patients with ACS who were admitted to Yokohama City University Medical Center and Kumamoto University Hospital, from January 1 2012, to December 31 2019. The inclusion criteria were age ≥18 years at enrollment, ACS diagnosis (unstable angina, non-ST elevation myocardial infarction [NSTEMI], or ST-elevation myocardial infarction [STEMI]) according to the Japanese guideline.^11^ The exclusion criteria were in-hospital death and discharge from hospitals with general wards stay ≤2 days, because it was difficult to evaluate defecation frequency. Clinical information was obtained from medical records.

This study was conducted in accordance with the guidelines of the Institutional Ethics Committees of two hospitals, which provided the ethical approvals (approval number 2766), and with the Declaration of Helsinki. Written informed consent was waived due to retrospective anonymized data collection. All patients enrolled had the opportunity to opt out through information posted on the webpages of both hospitals.

### Defecation frequency

Defecation data were collected from nursing records of individual patients. A defecation frequency of less than three times per week is commonly used to define chronic constipation. This criterion, however, cannot accurately assess an irregular defecation pattern, which includes both days with no defecation and days with frequent defecation. Thus, in addition to “average daily defecation frequency”, we comprehensively evaluated the following abnormal defecation patterns: (1) high frequency of days without defecation, (2) consecutive days without defecation, and (3) high-frequency defecation within a single day. Thus, we defined three indicators: “frequency of non-defecation days,” “consecutive non-defecation days,” and “maximum daily defecation frequency.” Days spent in the intensive care unit or high care unit were excluded from the calculation of defecation frequency. The “frequency of non-defecation days” was calculated as a ratio of days without defecation to the total days in a general ward. The examples of assessments for the indicators of defecation frequency can be found in **Figure S1**.

### Follow-up and definitions of adverse events

All patients were followed up for mortality, myocardial infarction, stroke, and heart failure hospitalization by reviewing their medical records and conducting phone calls with them or their families between January and August 2023. We also evaluated cause-specific mortality (cardiovascular, non-cardiovascular, and cancer mortality). The primary outcome was extended major adverse cardiovascular events (MACEs); a composite of the first occurrence of all-cause mortality, non-fatal myocardial infarction, non-fatal ischemic stroke, and non-fatal hemorrhagic stroke. The 3-point MACEs was a composite of cardiovascular mortality, non-fatal myocardial infarction, and non-fatal ischemic stroke.

### Statistical analyses

Statistical analyses were performed using JMP version 17.1.0 (SAS Institute Inc., Cary, NC). Variables are presented as a means ± standard deviations, medians (25th to 75th percentile), or numbers (%). Comparisons between the groups were performed using t-tests for parametric data and Wilcoxon test for non-parametric data. Categorical data were compared using Fisher’s exact test. Spearman’s correlation analysis was used to analyze the correlations between the indices. Receiver operating characteristic curve analysis was performed to evaluate the predictive ability of the four indicators of defecation frequency for primary outcomes. The cutoff values were determined using the maximum Youden’s index for the primary outcome. The Kaplan–Meier method was used for prognostic analysis, and the log-rank test was used to determine the significance of differences. Univariate and multivariate Cox regression analyses were used to assess independent prognostic capabilities. Age, sex, diabetes, prior myocardial infarction, chronic hemodialysis, diagnostic category of ACS, multivessel disease, Killip class, left ventricular ejection fraction, intra-aortic balloon pumping, coronary artery bypass grafting, venoarterial extracorporeal membrane oxygenation, β-blocker, calcium channel blocker, laxative, intestinal regulator, length of hospital stays, and food intake were used for adjustments in the multivariate Cox regression analyses and for conducting subgroup analyses. All statistical tests were two-sided, and statistical significance was set at *P*<0.05.

## Results

In total, 2119 patients were initially enrolled. **Figure S2** shows the patient enrollment process. After excluding patients with in-hospital deaths (n=93) and those with ≤2 days of general ward stay (n=77), 1949 patients remained (sex, 80.5% male; mean age, 67 years). The median (interquartile range [IQR]) length of stay in general wards was 12 (8–16) days, and the median (IQR) values for the four defecation indicators were 0.80 (0.57–1.11) times/day for the “average daily defecation frequency”, 33.3 (17.1–50.5) % for the “frequency of non-defecation days,” 2 (1–3) days for the “consecutive non-defecation days,” and 2 (1–3) times for the “maximum daily defecation frequency” (**Figure S3**). The cut-off values of four indicators of defecation for predicting the primary outcomes were 0.69 for the “average daily defecation frequency”, 33.5% for the “frequency of non-defecation days,” 6 days for the “consecutive non-defecation days,” and five times for the “maximum daily defecation frequency” (**Figure S4**).

### Clinical events during the follow-up

During the follow-up period (median, 48 months; interquartile range 30–74 months), 405 of 1949 (20.8%) patients developed the primary outcome. In total, 229 patients died (96 cardiovascular deaths and 133 non-cardiovascular deaths), and 278 patients experienced 3-point MACE. There were 142, 57, 28, and 113 patients with non-fatal myocardial infarction, nonfatal ischemic stroke, nonfatal hemorrhagic stroke, and hospitalized due to heart failure, respectively.

### Defecation frequency and the primary outcome

**Figure S5** illustrates the relationship between each indicator quartile and the primary outcome. **Figure 1** shows the univariate cumulative event rate analyses (Kaplan–Meier) and log-rank tests utilizing the cutoff values of the four defecation indicators, illustrating their association with the primary outcome. Notably, high “frequency of non-defecation days” was associated with events throughout the observation period, whereas high “maximum daily defecation frequency” suggested a particularly strong association with early events. The multivariate Cox hazard analysis in **Table 1** shows that high “frequency of non-defecation days” (adjusted hazard ratio [HR], 1.507; 95% confidence interval [CI], 1.191–1.907; *P*=0.0006) and high “maximum daily defecations frequency” (adjusted HR, 1.670; 95% CI, 1.203–2.317; *P*=0.002) have independent relationships with the primary outcome. Conversely, “consecutive non-defecation days” were not significantly associated with the events. When patients are divided into four groups using the cut-off values for “frequency of non-defecation days” and “maximum daily defecations frequency,” it becomes evident that risk stratification is possible from the early stages to the long term (**Figure 2**).

**Figure 1.**
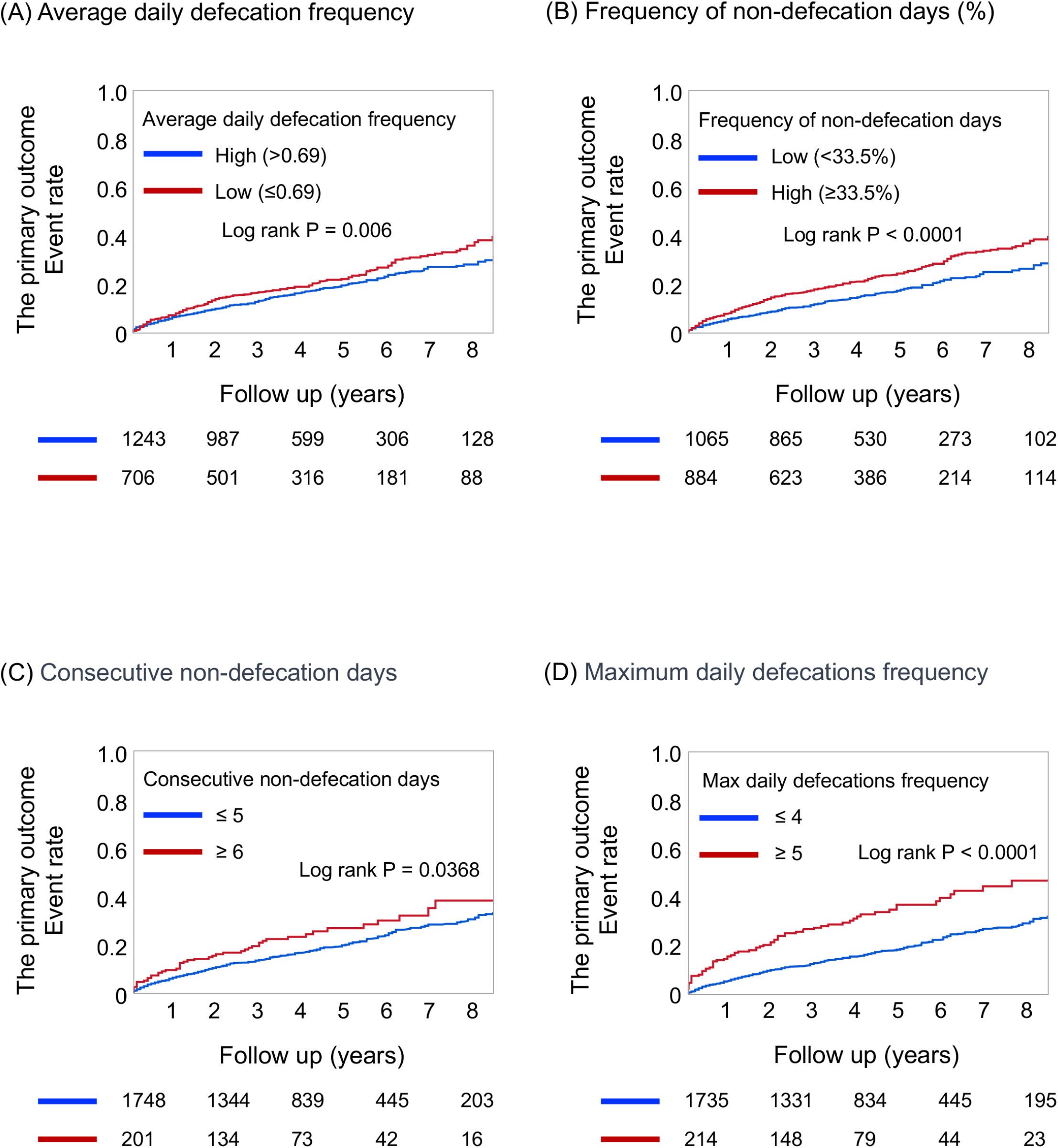
Kaplan–Meier curves for the composite primary outcome stratified by the four indicators of defecation frequency. The primary outcome was extended major adverse cardiovascular events (MACEs), which is a composite of the first occurrence of all-cause mortality, nonfatal myocardial infarction, nonfatal ischemic stroke, and nonfatal hemorrhagic stroke. The cut-off values of four indicators of defecation were 0.69 for the “average daily defecation frequency” (A), 33.5% for the “frequency of non-defecation days” (B), 6 days for the “consecutive non-defecation days” (C), and five times for the “maximum daily defecation frequency” (D).

**Figure 2.**
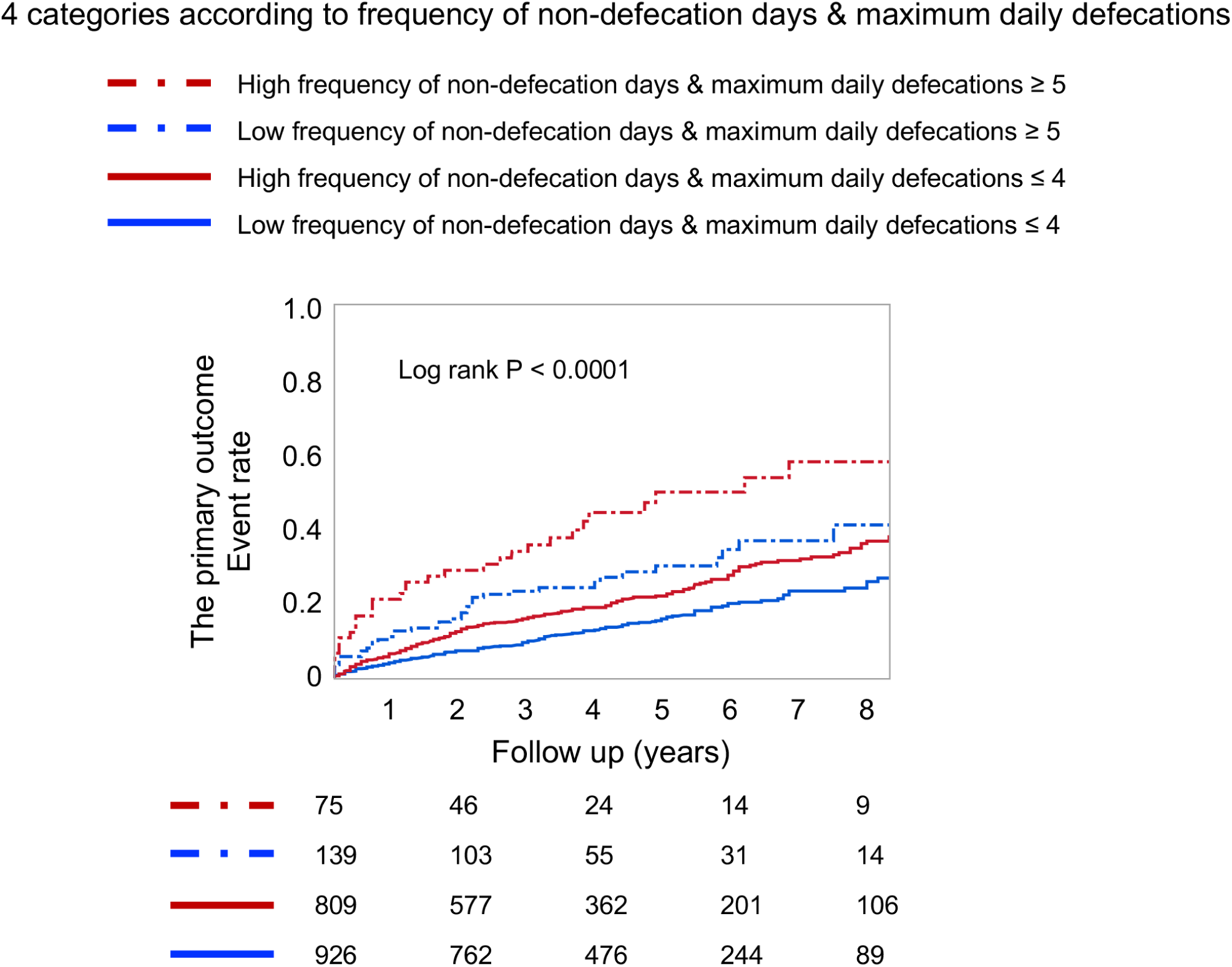
Kaplan–Meier curves for the composite primary outcome stratified by the “frequency of non-defecation days” and the “maximum daily defecations frequency.” The cut-off values of two indicators of defecation were 33.5% for the “frequency of non-defecation days,” and 5 times for the “maximum daily defecations frequency.”

**Table 1.**
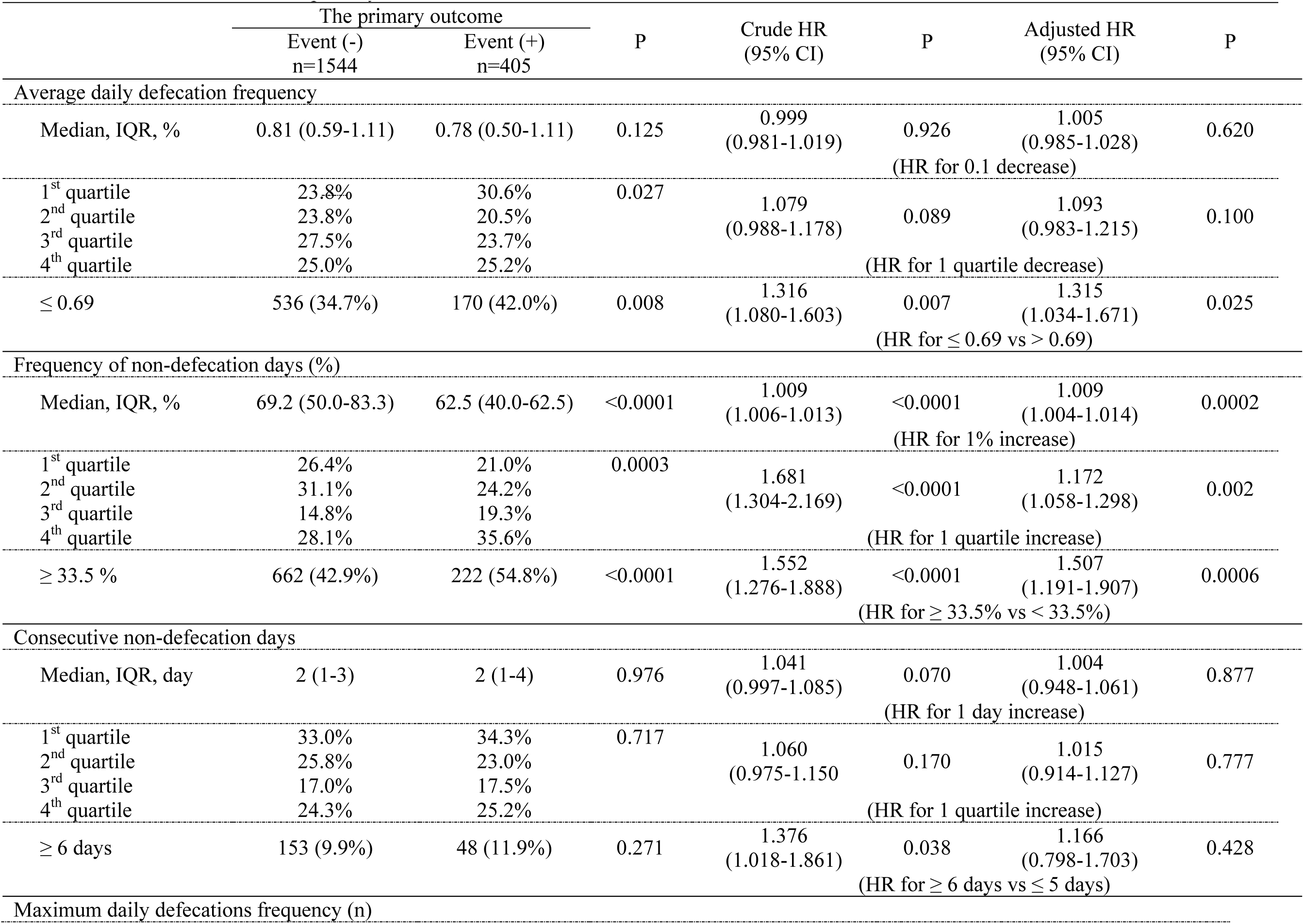
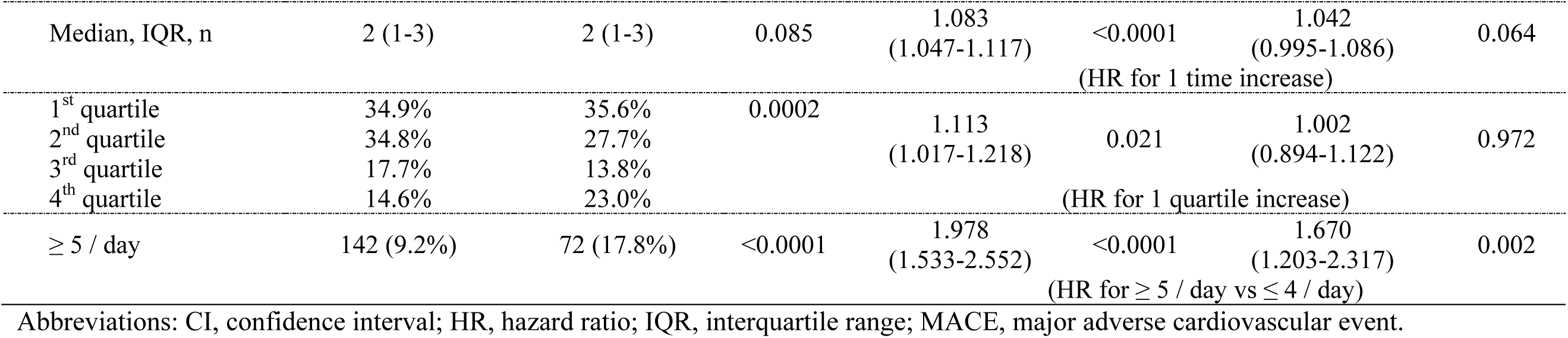
Defecation status and the primary outcome.

### Patients’ characteristics

The baseline patients’ demographics and clinical characteristics according to the “frequency of non-defecation days” and the “maximum daily defecations frequency” are listed in **Tables S1 and S2.** The high “frequency of non-defecation days” group had fewer cases of diabetes and STEMI compared to the low “frequency of non-defecation days” group. Additionally, the former group had more dialysis patients, a lower food intake, and shorter hospital stays. There was a modest negative correlation between the “frequency of non-defecation days” and the “maximum daily defecations frequency” (Spearman’s ρ=-0.33, *P*<0.0001) and the prevalence of high “maximum daily defecations frequency” was significantly lower in the group with high “frequency of non-defecation days” (**Table S1**). The high “maximum daily defecations frequency” group was characterized by older age, poorer kidney and heart function, and a higher proportion of severe cases requiring coronary artery bypass grafting, intra-aortic balloon pumping, or venoarterial extracorporeal membrane oxygenation compared to the low “maximum daily defecations frequency” group (**Table S2)**.

### Subgroup analysis

The subgroup analyses are shown in **Figures S6 and S7**. The higher risks in the high “frequency of non-defecation days” and high “maximum daily defecations frequency” groups for the primary outcome seemed to be consistent across all the subgroups.

### Details of clinical events

A high “frequency of non-defecation days” was significantly associated with 3-point MACE, all-cause death, cardiovascular death, non-cardiovascular death, non-fatal myocardial infarction, and hemorrhagic stroke (**Table 2**). The occurrence of ischemic stroke was numerically higher in the high than in the low “frequency of non-defecation days” group. Conversely, the high “maximum daily defecations frequency” group was significantly associated with 3-point MACE, non-cardiovascular death, cancer death, ischemic stroke, and heart failure hospitalization (**Table 3**).

**Table 2.** Adverse events and univariate Cox hazard analysis based on frequency of days without defecation.

|  | All<br>n=1949 | Frequency of non-defecation days |  | Crude HR | 95% CI | P |
| --- | --- | --- | --- | --- | --- | --- |
|  |  | low<br>< 33.5%<br>n=1065 | high<br>≥ 33.5%<br>n=884 |  |  |  |
| The primary outcome |  |  |  |  |  |  |
| Extended MACE<br>(all death, myocardial infarction, ischemic<br>and hemorrhagic stroke) | 405 (20.8%) | 183 (17.2%) | 222 (25.1%) | 1.552 | (1.276-1.888) | <0.0001 |
| 3-point MACE<br>(cardiovascular death, myocardial<br>infarction, and ischemic stroke) | 278 (14.3%) | 123 (11.6%) | 155 (17.5%) | 1.607 | (1.268-2.037) | <0.0001 |
| All death | 229 (11.8%) | 104 (9.8%) | 125 (14.1%) | 1.494 | (1.151-1.939) | 0.003 |
| Cardiovascular death | 96 (4.9%) | 42 (3.9%) | 54 (6.1%) | 1.593 | (1.064-2.386) | 0.024 |
| Non cardiovascular death | 133 (6.8%) | 62 (5.8%) | 71 (8.0%) | 1.425 | (1.013-2.005) | 0.042 |
| Cancer death | 63 (3.2%) | 34 (3.2%) | 29 (3.3%) | 1.075 | (0.655-1.766) | 0.774 |
| Non-fatal myocardial infarction | 142 (7.3%) | 60 (5.6%) | 82 (9.3%) | 1.738 | (1.245-2.426) | 0.001 |
| Non-fatal ischemic stroke | 57 (2.9%) | 26 (2.4%) | 31 (3.5%) | 1.510 | (0.896-2.543) | 0.122 |
| Non-fatal hemorrhagic stroke | 28 (1.4%) | 8 (0.8%) | 20 (2.3%) | 3.128 | (1.375-7.114) | 0.007 |
| Heart failure hospitalization | 113 (5.8%) | 62 (5.8%) | 51 (5.8%) | 1.072 | (0.740-1.553) | 0.713 |

1 Abbreviations: CI, confidence interval; HR, hazard ratio; MACE, major adverse cardiovascular event.

**Table 3.**
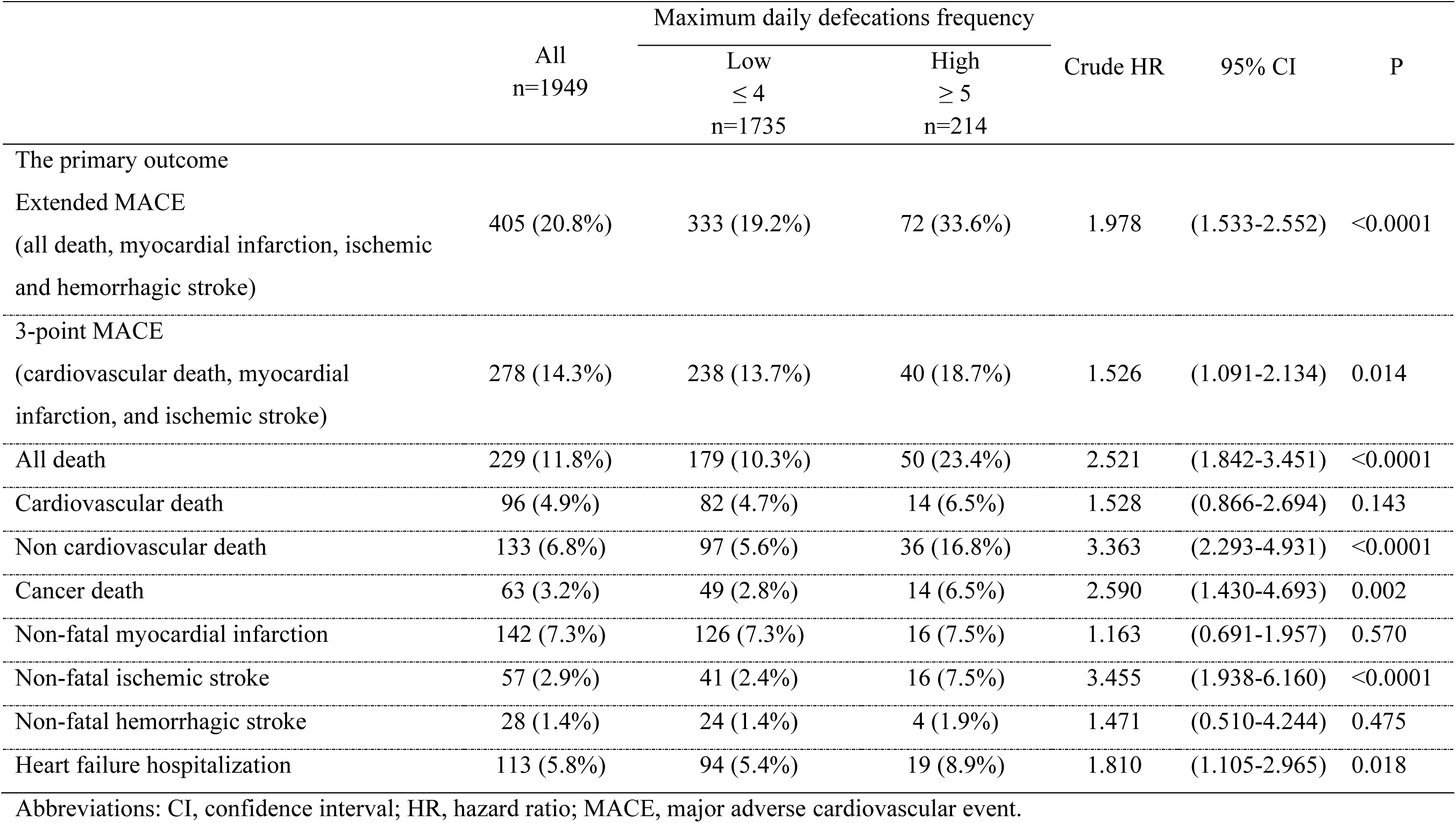
Adverse events and univariate Cox hazard analysis based on maximum daily defecations frequency.

The high “frequency of non-defecation days” and high “maximum daily defecations frequency” groups exhibited differences in the breakdown of associated events and variations in timing, spanning both early and long-term outcomes. Consequently, a defecation pattern characterized by infrequent defecations on certain days but high-frequency defecations on other days was prognostically unfavorable (**Figure 2**).

## Discussion

**Figure 3** illustrates the summary of this study. High “frequency of non-defecation days” and high “maximum daily defecations frequency” during their general wards stay were associated with a heightened risk of future cardiovascular events. These associations remained significant even after adjusting for factors, such as clinical presentation and severity of ACS, age, sex, dietary intake, length of hospital stay, and laxative use. These two indicators exhibit distinct characteristics; a high “frequency of non-defecation days” is associated with the occurrence of cardiovascular mortality, all-cause mortality, myocardial infarction, and cerebral hemorrhage over an extended period. Conversely, a high “maximum daily defecation frequency” is linked to cancer-related mortality, non-cardiovascular mortality, cerebral infarction, and heart failure, especially in the early phase. To our knowledge, this study is the first to report the defecation frequency during the acute phase in patients with ACS and utilize reliable records of defecation during hospitalization rather than relying on survey-based assessments.

**Figure 3.**
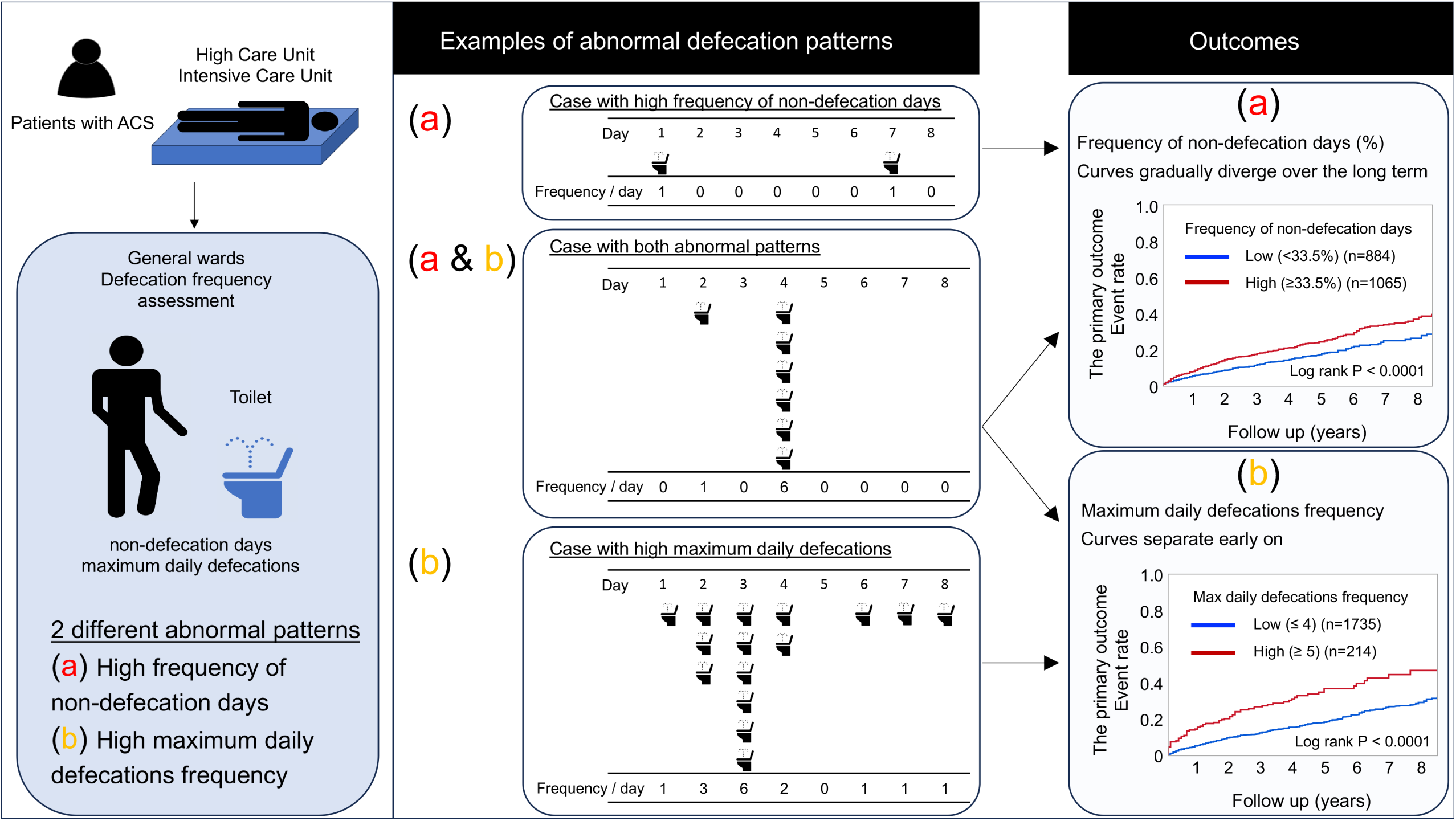
Summary of the study. In this study, defecation patterns among patients with acute coronary syndrome (ACS) who survived to discharge were investigated using three indicators: “frequency of non-defecation days,” “consecutive non-defecation days,” and “maximum daily defecation frequency,” in addition to “average daily defecation frequency”. Both increased frequency of non-defecation days and high-frequency defecations within a single day were independently associated with an increased risk of future cardiovascular events, controlling for various confounding factors including age, severity of ACS, medications, and dietary intake. Further research is needed to explore the mechanisms and potential therapeutic interventions.

The observed association between high frequency of in-hospital non-defecation days and subsequent cardiovascular events in patients with ACS in this study aligns with prior research findings indicating that “chronic constipation is linked to an adverse cardiovascular prognosis.”^6–8^ One notable difference between previous studies and this study is the result of heart failure outcome. Previously,^6^ a significant association between constipation and heart failure was reported. However, in this study, no significant relationship was observed between the high “frequency of non-defecation days” and heart failure events. In the present study population, advanced age, female sex, Killip class ≥2, and reduced left ventricular ejection fraction were identified as factors associated with heart failure events (data not shown). However, when comparing the high and low “frequency of non-defecation days” groups, age, sex, and Killip class were similar, while the left ventricular ejection fraction was unexpectedly higher. Patients’ backgrounds may have introduced confounding factors that hindered the detection of an apparent association between a low defecation frequency and heart failure events. Conversely, high “maximum daily defecations frequency” was initially associated with heart failure events, but upon adjustment for the aforementioned factors, this relationship lost its statistical significance (data not shown). Hence, further research and analysis are imperative to achieve a more thorough understanding of the potential link between defecation frequency and heart failure events, while meticulously considering these potential confounding factors.

The high “frequency of non-defecation days” group had a higher proportion of patients who had previously experienced myocardial infarction or developed renal failure requiring chronic dialysis. By including ACS events for the inclusion of this study, it can be inferred that patients with impaired gut function had a higher frequency of recurrent cardiovascular events. These results suggest a potential association between decreased gut function and cardiovascular diseases. Furthermore, the high “frequency of non-defecation days” group exhibited a lower proportion of STEMI cases and a higher proportion of NSTEMI cases. Within the current study population, patients with a previous history of myocardial infarction demonstrated a significantly higher incidence of NSTEMI compared to those without (58.3% vs. 34.5%, *P*<0.0001). Similarly, patients undergoing chronic hemodialysis had a significantly higher incidence of NSTEMI than those who did not (78.5% vs. 35.1%; *P*<0.0001), suggesting that the differential frequencies of STEMI and NSTEMI in the two groups stratified by the “frequency of non-defecation days” can be attributed, at least in part, to these factors. Notably, the association between high “frequency of non-defecation days” and unfavorable cardiovascular prognosis remained consistent in the subgroup analyses stratified by STEMI/NSTEMI, prior history of myocardial infarction, and chronic hemodialysis.

In this study, the association between high “frequency of non-defecation days” and cardiovascular events remained independent of various confounders. Consequently, while acute stress, impaired cardiac function, and renal dysfunction may contribute to decreased defecation frequency, it is suggested that the increased “frequency of non-defecation days” may also be linked to cardiovascular events through distinct mechanisms. In patients with severe ACS, characterized by cardiac dysfunction, renal impairment, heart failure, and those requiring treatments, such as coronary artery bypass grafting or mechanical circulatory support, a reduction in defecation frequency was not observed; rather, a higher “maximum daily defecations frequency” was noted. This observation may be associated with the higher incidence of early events in the high “maximum daily defecation frequency” group in this work. The two indicators used to evaluate the defecation frequency may reflect different aspects of patient prognosis. There was a small group of patients (n=75, 3.9%) whose overall hospitalization period had a low number of days with defecation, but who exhibited a high maximum daily defecation frequency on specific days. The patients in this group had the worst prognoses, which cannot be detected by the “average daily defecation frequency”.

As this study focused on the period when patients were hospitalized in general wards, receiving dietary intake, and undergoing rehabilitation, the defecation frequency during this timeframe can be considered indicative of gut function. Notably, the high “frequency of non-defecation days” displayed a robust association with cardiovascular mortality and myocardial infarction. A long gut transit time has been reported to be associated with gut microbial diversity, composition, and metabolism, leading to chronic inflammatory responses that may elevate the risk of cardiovascular diseases.^12–16^ Conversely, certain types of gut microbial dysbiosis can dampen gut motility, establishing a bidirectional relationship between the gut microbiota and gut transit time.^17^ Moreover, other types of gut function abnormalities, such as irritable bowel syndrome, also influence nutrient absorption and gut microbiota.^18^ Nonetheless, the association between cardiovascular events and defecation frequency remains under investigation, and the precise mechanisms and causal relationships remain unclear. Investigations into why a high “maximum daily defecations frequency” during acute illness is associated with subsequent cardiovascular events have not been conducted. Additionally, this study was retrospective in nature, with a focus solely on defecation frequency for assessment and analysis. Precise examinations of the gut transit time using methods, such as radio-opaque markers, scintigraphy with radioactive isotopes, and wireless capsule techniques for measuring intraluminal pressure, temperature, and pH, have not been performed. Nevertheless, the study was simple and non-invasive, allowing evaluation in a wide range of individuals. In contrast to previous studies that relied on questionnaire surveys, the data on defecation frequency were meticulously recorded by nurses during hospitalization, ensuring high reliability. Our findings may reflect an aspect within the broader pathological concept of impaired gut function, which could be described as “gut frailty.” However, the evaluation of gut function was not conducted precisely, and variables, such as gut microbiota, metabolites originating from the gut microbiota, leaky gut syndrome, and incretin secretion, were not assessed. Therefore, the specific mechanisms underlying these results remain unclear. Although numerous reports and research advancements have explored the relationship between physical frailty and cardiovascular diseases, limited research has focused on the connection between gut function and cardiovascular diseases. Further investigations are necessary to determine whether interventions aimed at improving gut function, such as increased dietary fiber intake, judicious use of laxatives, or adoption of moderate exercise routines, can enhance cardiovascular prognosis.

### Limitations

This study was a retrospective, observational, two-center investigation; thus, it has inherent limitations, including a relatively small sample size and the inability to establish causality. Given the absence of data on defecation frequency before and after hospitalization, it remains uncertain whether the observed indicators of defecation frequency are solely acute-phase phenomena or chronic conditions. Nevertheless, the Kaplan–Meier curves suggest that a higher frequency of non-defecation days during hospitalization may be linked to long-term event occurrence. Only defecation frequency was available, and assessments of stool characteristics using tools, such as the Bristol Stool Chart, were not conducted. Furthermore, this study included only Japanese patients with ACS; therefore, the findings cannot be directly extrapolated to other populations.

## Conclusions

Among patients discharged after hospitalization for ACS, high “frequency of non-defecation days” and high “maximum daily defecations frequency” during general wards stay were associated with an increased risk of future cardiovascular events. When defecation occurs on ≤2 out of 3 days, the long-term risk of future cardiovascular events increases by approximately 50%. Conversely, when there was a day with five or more defecations, the risk of early events was heightened. Further research is required to gain a deeper understanding of the underlying mechanisms. These additional investigations will yield valuable insights into the association between defecation frequency and cardiovascular events.

## Clinical Perspectives

### COMPETENCY IN PATIENT CARE AND PROCEDURAL SKILLS

In patients with acute coronary syndrome, abnormal defecation patterns during general wards stay are associated with major adverse events after discharge.

### TRANSLATIONAL OUTLOOK

Further research is needed to explore the mechanisms and potential therapeutic interventions.

## Data Availability

All data is available upon approval. It will be necessary to provide a detailed protocol for the proposed study, to provide the approval of an ethics committee, to supply information about the funding and resources one has to carry out the study, and to consider inviting the original authors to participate in the re-analysis.

## Acknowledgements

We thank Mayu Aoki and Ryuta Shirahama at Yokohama City University Medical Center and Kei Nakashima at Kumamoto University Hospital for data management.

## Data availability statement

The data produced and analysed in this study are available from the corresponding author upon reasonable request.

## Study Setting

Yokohama City University Medical Center and Kumamoto University Hospital.

## Abbreviations

ACS: acute coronary syndrome
CI: confidence interval
HR: hazard ratio
MACE: major adverse cardiovascular events
NSTEMI: non-ST elevation myocardial infarction
STEMI: ST-elevation myocardial infarction

